# Comparing Cardiac Genetic Testing Pathways: Impacts on Access, Informed Choice, and Decisional Satisfaction

**DOI:** 10.64898/2026.04.03.26350137

**Authors:** Susan Christian, Talia C. Belcher, Marc Benoit, Alicia Chan, Tara Dzwiniel, Erkan Ilhan, Shailly Jain-Ghai, Karen Katchmer, Omid Kiamanesh, Margaret Lilley, Julien Marcadier, Skylar Moreau, Andrew Muranyi, Ashley J. Nicolas, Priyana Sharma, Xi Zhao, Cathleen Huculak

## Abstract

**Background:** Mainstreaming genetic testing has emerged as a strategy to improve access and reduce wait times for patients who may benefit from genetic testing. Ensuring patients fully grasp the implications of testing when formal genetic counselling is not provided, remains a focus for ongoing research.

**Methods:** Patients diagnosed with hypertrophic or dilated cardiomyopathy were offered genetic testing between September 2024 and September 2025 through either the mainstreaming model conducted in cardiology clinics or a referral to Medical Genetics where patients attended an online webinar or a one-on-one genetic counselling appointment. Uptake of testing, time to testing, informed choice and patient satisfaction were evaluated.

**Results:** Among patients offered genetic testing, uptake was higher in the mainstreaming pathway (82%) compared with a referral to Medical Genetics (69%). The difference in access was predominately due to patients not following through with their Genetics referral. Mainstreaming reduced wait times where patients referred to Genetics waited a median of 94-185 additional days to be offered genetic testing. Despite improved access, only 62% of mainstreamed patients were considered informed, compared to 91% of patients that attended a patient webinar through Medical Genetics (p < 0.01). Satisfaction with decision-making was high across both pathways.

**Conclusion:** Integrating genetic testing into cardiology practices increased access and reduced wait times; however, patients demonstrated significantly lower rates of informed decision making compared to those who attended a patient webinar offered through Medical Genetics. These findings highlight the importance of structured education to support informed decision-making within mainstreaming pathways.

## Introduction

Cardiac genetic testing is recommended as part of diagnostic investigations for cardiomyopathies and has shown promise in guiding management for individuals carrying disease causing variants in specific genes.^1^ It also plays a pivotal role in identifying at-risk family members who may otherwise not be recognized for cardiac screening. Due to increasing demand and limited genetic counselling resources, several service delivery models have been implemented to improve access. These include in-person group sessions (pre-COVID), live patient webinars, and embedding genetic counsellors into multidisciplinary clinics.^2–6^ Despite these efforts, referral volumes continue to rise and wait times remain unacceptable at many centers.

To improve timely access to testing, alternative models such as mainstreaming have emerged as promising solutions. Mainstreaming is defined as “shifting all or part of the clinical genetic testing process to clinicians outside of the genetics service to facilitate patient access to genetic testing.”^7^ Most mainstreaming models that have been implemented to date involve nongenetic clinicians performing assessment activities and pre-test activities but vary with regards to post-test activities such as result disclosure, clinical correlation, post-disclosure navigation, segregation analysis, cascade testing in at-risk family members, and reproductive counselling.^7^ Mainstreaming aims to increase access, reduce wait times, and optimize the use of limited genetic counselling resources by shifting pre-test activities to non-genetic clinicians, allowing Medical Genetics to focus on post-test counselling, segregation analysis, and cascade predictive testing. However, it requires careful implementation to ensure patients receive adequate information to make informed decisions and understand their results.

In oncology, mainstreaming has been extensively studied, particularly in the context of hereditary breast and ovarian cancer. Research has shown that mainstreaming increases the uptake of genetic testing, reduces wait times, and lowers healthcare costs.^8^ Patient satisfaction with this model of care is generally high, and levels of anxiety or decisional regret are comparable to those seen in traditional counselling pathways.^9^ Moreover, mainstreaming has been shown to reduce disparities in access to testing, specifically among underserved populations.^10^

In cardiology, the application of mainstreaming is more recent, less widespread, and not as well studied. Despite guidelines supporting genetic testing for hypertrophic cardiomyopathy (HCM) and dilated cardiomyopathy (DCM), testing rates remain low. ^11–16^ Emerging studies suggest that cardiologists are interested in incorporating genetic testing into their practice but face barriers such as limited genetics training, lack of infrastructure, and concerns about patient understanding.^17^ Robust data on patient outcomes and system-level impacts in cardiology and medical genetics are limited.

Across medical specialties, a central goal is ensuring patients understand key concepts related to genetic testing and make informed decisions. This means providing clear, relevant information about the benefits, limitations, and possible outcomes of testing, for the individual and their family. It also involves supporting patients in making choices that reflect their personal values and preferences. Although informed choice has not specifically been evaluated for mainstreaming models, it is generally felt that with tools, such as checklists, educational videos, and written materials, non-genetic clinicians can effectively convey the essential components of genetic testing. Frameworks like CADRe <u>(Consent & Disclosure Recommendations) - ClinGen | Clinical Genome Resource</u>) have been developed to help clinicians deliver concise pre-test information. However, ensuring patients fully grasp the implications of testing when formal genetic counselling is not provided, remains a focus for ongoing research.

This study aimed to evaluate the impact of a mainstreaming approach to cardiac genetic testing in Alberta. The study assessed access to genetic testing, patient experience, patients’ ability to make an informed choice, and patient satisfaction with their decision.

## Methods

In Alberta, Canada, genetic testing for HCM and DCM has been restricted to medical genetic specialists. Patients are referred to one of the two tertiary care centres, located in Edmonton (Medical Genetic Clinic) and Calgary (Clinical Genetics). They are given the option to attend a provincial HCM or DCM patient webinar, followed by a short phone call with a genetic counsellor; or wait for a one-on-one appointment with a genetic counsellor. To address the increasing wait times to see a genetic counsellor, a real- world implementation study was performed from September 9, 2024 to September 9, 2025. In this model, non-genetic specialists initiated genetic testing without a prior referral to genetic counselling.

### Real-world Implementation Study

Six hospital and community-based cardiac specialists from across Alberta were invited to participate in a one-year real-world implementation study, trialing a mainstreaming model. The cardiac specialists were identified based on a high volume of prior referrals to Medical Genetics. At the start of the study, participating cardiac specialists received a short virtual presentation from the cardiac genetic counselling team outlining the logistics of the study and key information that should be reviewed with patients when offering genetic testing. Key information included the benefits of genetic testing (providing an opportunity for cascade genetic testing), importance of communicating genetic results with relatives, limitations of genetic testing, and possibility of an unclear result. Additional important concepts included autosomal dominant inheritance, associated risks (sudden cardiac arrest), incomplete penetrance, and variable expression. Cardiac specialists were given a toolkit that included a checklist of the key information to discuss with patients, a list of frequently asked questions, written patient educational material, and a link to a patient-facing video (Supplementary Material). The resources were designed to support informed choice in the absence of formal genetic counselling.

Cardiac specialists were advised to refer patients with a complex presentation, complicated family history, or greater counselling needs to Medical Genetics. Upon receipt of genetic test results, cardiac specialists were asked to disclose the findings to their patients. If deemed appropriate, those with negative results received a family letter outlining cardiac screening recommendations for first-degree relatives. Patients with positive or uncertain results were referred to Medical Genetics for post-test genetic counselling.

### Study group

Eligible patients were 18 years or older and had a clinical diagnosis of HCM (left ventricular hypertrophy ≥15 mm) or DCM (dilated left ventricle and left ventricular ejection fraction <50%). Cardiac specialists discussed the option of genetic testing with patients and provided them with a test requisition for a cardiac gene panel. Patients who were seen in an outpatient setting and were fluent in reading English were also invited to complete an online survey about their experience. Patients who were offered genetic testing while hospitalized were not invited to complete the survey, due to the potentially stressful nature of their clinical circumstances.

### Comparison groups

Access (uptake of genetic testing and time to genetic testing) was compared between patients offered genetic testing through the mainstreaming model and patients referred to the Edmonton Medical Genetics Clinics. This included patients that attended a webinar, patients that requested a one-on-one genetic counselling appointment, patients that declined a Medical Genetics appointment, and those that were lost to follow-up after three or more attempts to schedule an appointment. Access was not evaluated for patients referred to the Calgary Clinical Genetics Unit as not all outcome data was available to the research team..

To evaluate patient experience, patient satisfaction with their decision, and patients’ ability to make an informed choice, survey responses were compared between patients seen through the mainstreaming models and patients seen through the existing provincial HCM/DCM webinar-based model.^18^ Patients meeting the same inclusion criteria and who attended a webinar through the Edmonton or Calgary Genetics Clinics were also invited to complete a patient survey. Disease presentation and severity of webinar participants were expected to be comparable to those seen through the mainstreaming model.

### Outcomes

#### 1. Access to genetic testing

##### Uptake of genetic testing

We compared the proportion of patients who had genetic testing completed through the mainstreaming model with those referred to the Edmonton Medical Genetics Clinic. Uptake of genetic testing was defined as the proportion of patients within each group that proceeded to have a sample collected by December 31, 2025. Reason for not having a sample collected included patients declining genetic counselling, declining genetic testing, not having a sample collected, or being lost to follow up up after three or more attempts to schedule an appointment. Age, gender, and diagnosis were extracted from the patient’s electronic record. Uptake of genetic testing was compared across patient demographics using logistic regression. All statistical analyses were conducted using Stata software (StataCorp LP, College Station, TX

##### Time to genetic testing being offered

Time to being offered genetic testing in the mainstreaming group was considered immediate (0 days). For patients referred to the Edmonton Medical Genetics Clinic, the median time (IQR) to being offered genetic testing was calculated based on referral date and date seen (in days).

#### 2. Patient Experience, Satisfaction with Decision, and Informed Choice

Eligible patients who were offered genetic testing as an outpatient through the mainstreaming model and those that attended an HCM/DCM webinar were invited to complete an online survey shortly after of being seen. The survey included demographic questions (age, gender, ancestry/race, education, and diagnosis), questions about their experience, the Satisfaction with Decision scale and Cardiac Multidimensional Model of Informed Choice (MMIC) scale (Supplementary Material). Linear and logistic regression were performed to evaluate associations between satisfaction with decision and informed choice based on demographic variables. Chi squared analysis was used to compare knowledge and informed choice based on the model of care. A p-value of <0.05 was defined as statistically significant.

##### Patient experience

Participants in both groups were asked if they were (1) provided sufficient time to talk about whether they wanted genetic testing, (2) informed when results would be available, and (3) informed how they would receive the result. Participants in the mainstreaming group were also asked if they were (1) given a patient information sheet about genetic testing, (2) told that they could talk with a genetic counsellor before deciding about genetic testing, and (3) informed that they would be referred to a genetic counsellor if a genetic cause was found for their heart condition.

##### Satisfaction with Decision

The Satisfaction with Decision Scale is a six-item validated tool that measures individual satisfaction about a health-related decision.^19^ It includes a 5-point Likert scale and is commonly used in genetic counselling research to assess decision quality and confidence. Each item was rated on a scale from 1 (strongly disagree) to 5 (strongly agree), with higher scores indicating greater satisfaction. Total scores were calculated by summing responses across completed items. If a participant skipped a question, the total score was adjusted by reducing the denominator accordingly to reflect the number of items answered. Descriptive statistics were used to describe overall satisfaction with decision.

##### Informed Choice

The Cardiac MMIC Scale is a novel tool developed to assess whether patients with inherited cardiac conditions (i.e., DCM and HCM) are making informed decisions about genetic testing.^20^ The scale includes eight true/false knowledge questions based on core concepts routinely addressed in cardiac genetic counselling. Participants were also given the option to respond with “I don’t know” to each knowledge question, which was treated as an incorrect response. This allowed for a more nuanced assessment of patient understanding. The scale also includes five questions about the patient’s attitude toward genetic testing and one question asking if they intended to have genetic testing. Informed choice was defined as having adequate knowledge (a score of ≥75%) and an attitude that aligned with the participant’s intention to undergo genetic testing. Descriptive statistics were used to categorize participants as informed or not informed.

Ethics approval was obtained through the Research Ethics Board at the University of Alberta (Pro00140655 and Pro00161643) and the Conjoint Health Research Ethics Board at the University of Calgary (pSite-24-0036).

## RESULTS

### 1. Access to genetic testing

#### Uptake of genetic testing

Between September 9, 2024 and September 9, 2025, 91 patients with DCM or HCM were offered cardiac genetic testing in an outpatient setting through the mainstreaming model (Figure 1). An additional 25 in-patients were offered genetic testing through mainstreaming. Of these patients, 95 had a sample collected (25/25 in-patients and 70/91 outpatients). The cardiac specialists reported that a small number of patients declined genetic testing upfront (specific numbers not recorded). Therefore, the uptake of genetic testing through the mainstreaming model was 82% (95/116).

**Figure 1:**
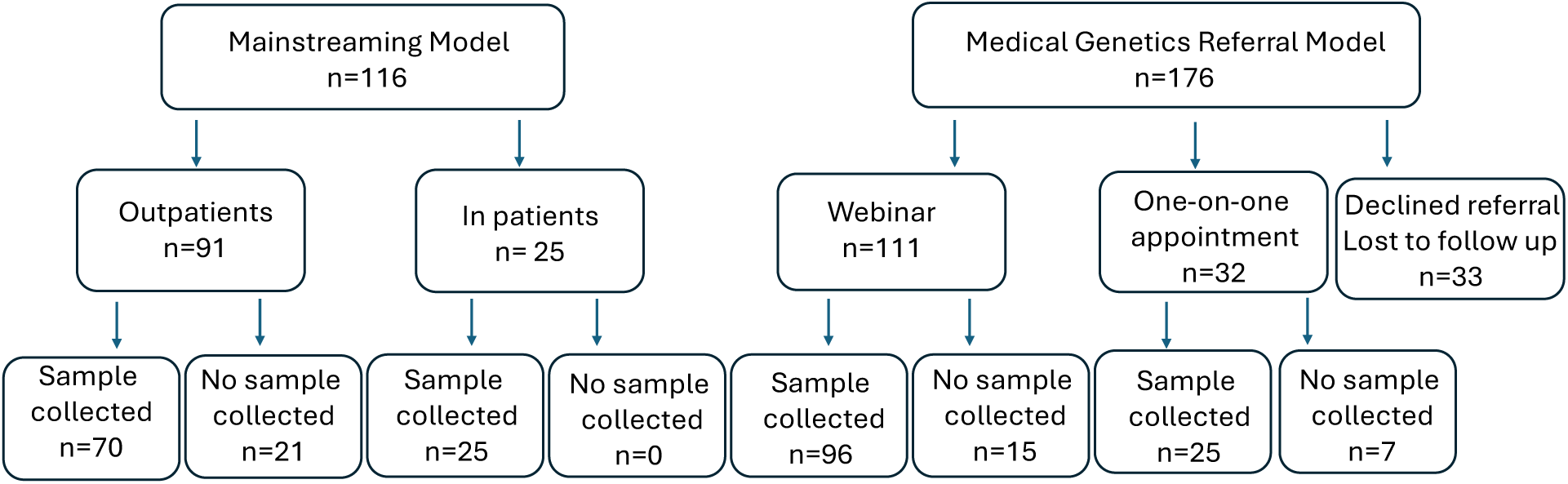
Uptake of genetic testing in mainstreaming model compared to medical genetics referral model.

During this same time frame, 176 patients with a diagnosis of DCM or HCM were referred to the Edmonton Medical Genetics Clinic for genetic counselling (Figure 1). Of these, 111 were seen through the webinar pathway with three declining genetic testing upfront, and 96 proceeding with sample collection. An additional 32 patients were seen through one-on-one genetic counselling appointments, resulting in 25 samples being collected. All additional patients were either lost to follow-up (n=20) (i.e. unable to schedule an appointment after 3 attempts) or declined the referral (n=12), and one patient died before being seen. Therefore, the uptake of genetic testing for patients referred to Medical Genetics was 69% (121/176).

Demographic data for patients seen in each group is shown in Table 1. Uptake of genetic testing was higher in woman compared to men in the webinar group (OR= 2.24; 95% CI [1.05, 4.79]; p=0.05) and although a similar trend was seen in the mainstreaming group the difference was not significant (OR= 1.74; 95% CI [0.62, 4.88]; p=0.29) (Table 2). Uptake did not differ significantly based on age or diagnosis in either group.

**Table 1:**
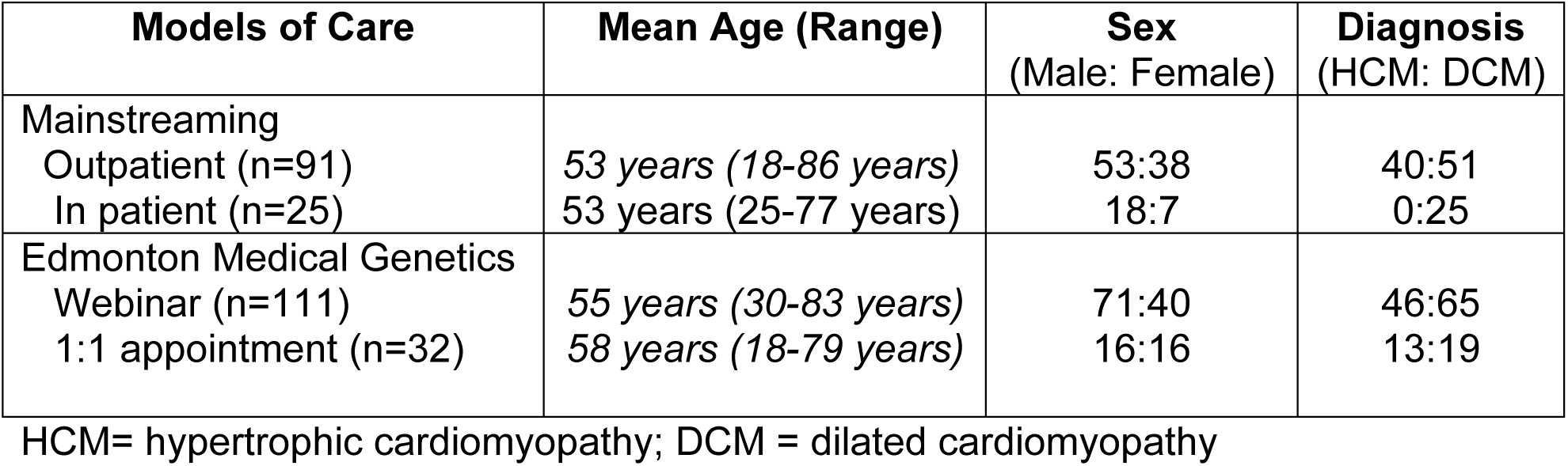
Demographics of patients offered genetic testing through mainstreaming and the Edmonton Medical Genetics Clinic.

**Table 2.** Uptake based on demographic variables.

|  | Medical Genetic |  | Mainstreaming |  |
| --- | --- | --- | --- | --- |
|  | Odds Ratio [95% CI] | p value | Odds ratio [95% CI] | p value |
| DCM vs HCM | 0.96 [0.52, 1.78] | 0.90 | 1.16 [0.44, 3.07] | 0.77 |
| Increasing Age | 1.02 [0.99, 1.04] | 0.21 | 1.03 [1.00, 1.07] | 0.08 |
| Female vs Male | 2.24 [1.05, 4.79] | 0.04 | 1.74 [0.62, 4.88] | 0.29 |

#### Time to genetic testing

The median time for patients referred to the Edmonton Medical Genetics Clinic to attend the webinar was 94 days (IQR 60-201, range 13–675). Some delays to be seen were patient related (i.e. no-showing to previous webinars or scheduling conflicts). The median time to be seen for a one-on-one genetic counselling appointment was 185 days (IQR 118-224, range 32-400 days).

### 2. Patient experience, satisfaction with decision, and informed choice

In the mainstreaming group, 91 outpatients were offered genetic testing, of whom 84 were invited to complete the patient survey (Figure 2). Of these, 81 consented to the survey, while 3 declined. In the provincial webinar group, 228 patients were offered genetic testing, 206 were invited to participate, and 186 consented to the survey, and 20 declined. Survey completion rates were 75% (61/81) in the mainstreaming group and 65% (121/186) in the webinar group. Demographic data for survey participants is shown in Table 3.

**Figure 2:**
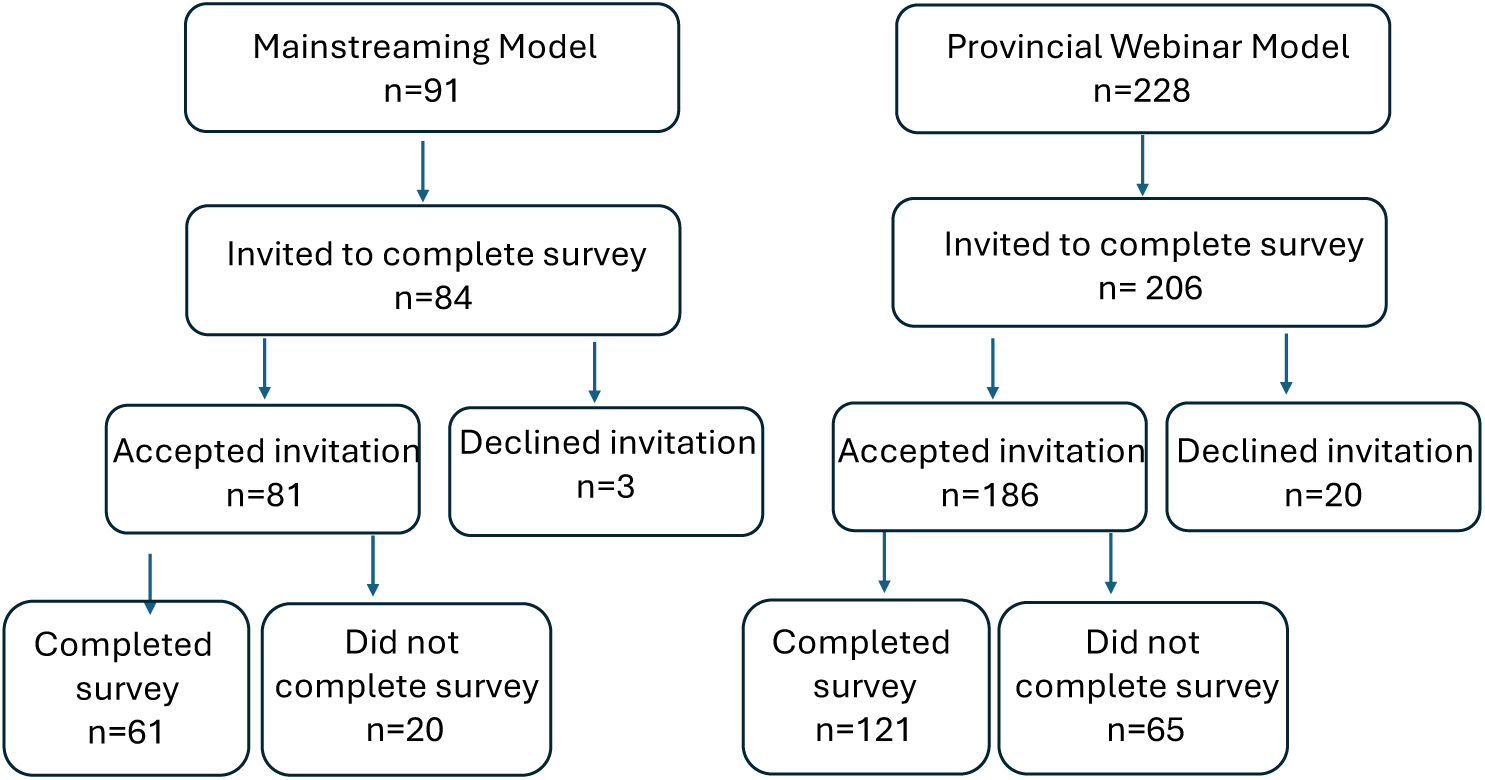
Survey completion in mainstreaming model compared to provincial webinar model.

**Table 3:**
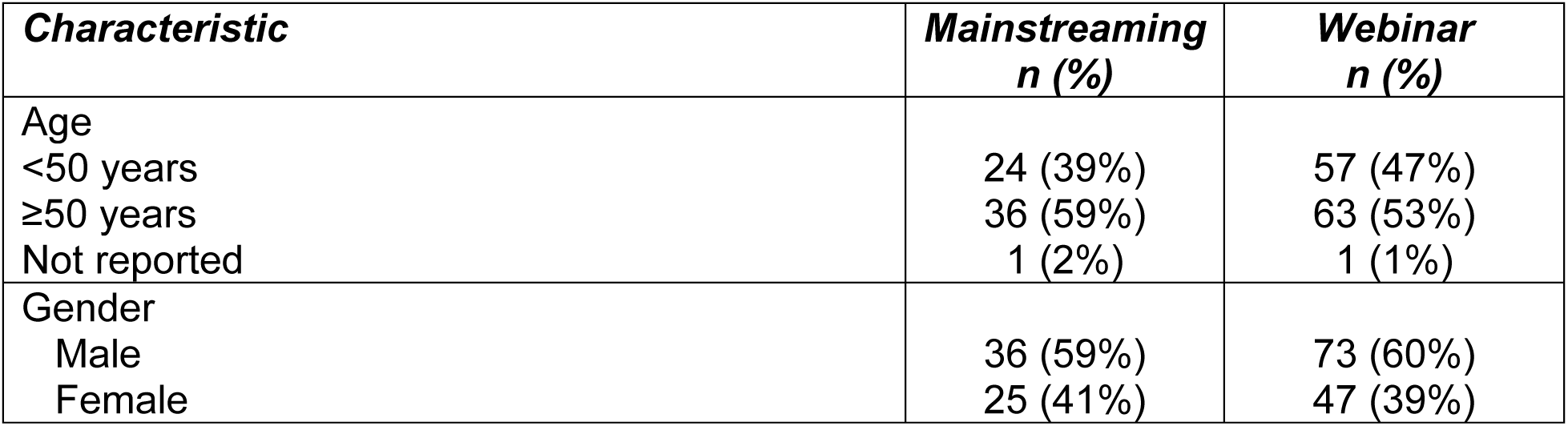
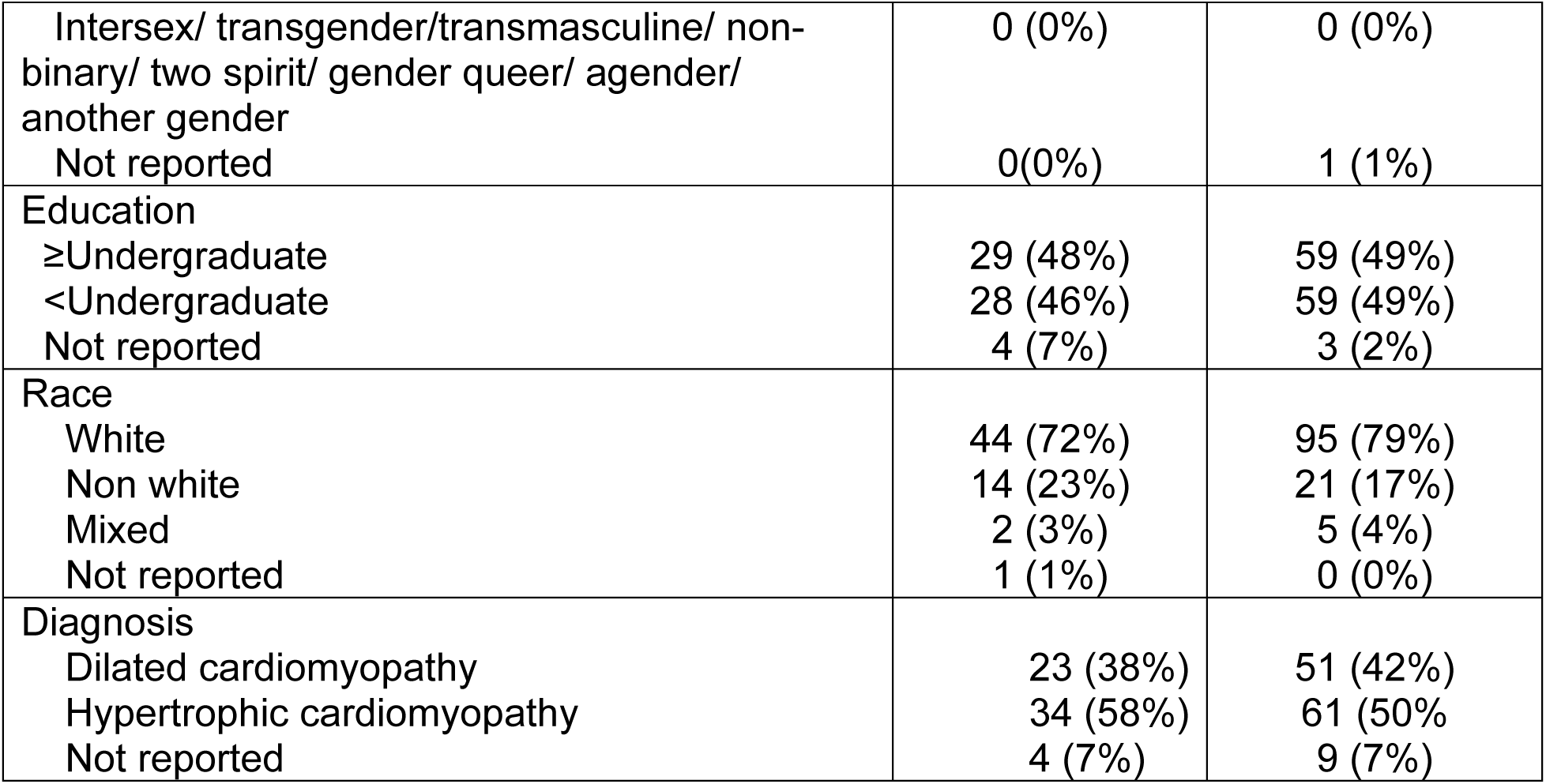
Demographics of participants that completed the patient survey.

| Characteristic | Mainstreaming<br>n (%) | Webinar<br>n (%) |
| --- | --- | --- |
| Age |  |  |
| <50 years | 24 (39%) | 57 (47%) |
| ≥50 years | 36 (59%) | 63 (53%) |
| Not reported | 1 (2%) | 1 (1%) |
| Gender |  |  |
| Male | 36 (59%) | 73 (60%) |
| Female | 25 (41%) | 47 (39%) |
| Intersex/ transgender/transmasculine/ non-binary/ two spirit/ gender queer/ agender/ another gender | 0 (0%) | 0 (0%) |
| Not reported | 0(0%) | 1 (1%) |
| Education |  |  |
| ≥Undergraduate | 29 (48%) | 59 (49%) |
| <Undergraduate | 28 (46%) | 59 (49%) |
| Not reported | 4 (7%) | 3 (2%) |
| Race |  |  |
| White | 44 (72%) | 95 (79%) |
| Non white | 14 (23%) | 21 (17%) |
| Mixed | 2 (3%) | 5 (4%) |
| Not reported | 1 (1%) | 0 (0%) |
| Diagnosis |  |  |
| Dilated cardiomyopathy | 23 (38%) | 51 (42%) |
| Hypertrophic cardiomyopathy | 34 (58%) | 61 (50%) |
| Not reported | 4 (7%) | 9 (7%) |

#### Patient experience

Within the mainstreaming group, 97% (58/60) of participants indicated that they were given sufficient time to talk about whether they wanted genetic testing, 61% (37/61) indicated that they were informed when results would be available, and 55% (33/60) stated that they were informed how they would receive the result. Further, 59% (35/61) of participants in the mainstreaming group were given the patient information sheet about genetic testing, 78% (46/59) were told that they could talk with a genetic counsellor before deciding about genetic testing, and 74% (45/61) were told that they would be referred to a genetics counsellor if a genetic cause was found for their heart condition. In comparison, in the webinar group, 99% (120/121) of participants indicated that they were given sufficient time to talk about whether they wanted genetic testing, 95% (115/121) indicated that they were informed when results would be available, and 95% (115/121) stated that they were informed how they would receive the result.

#### Satisfaction with Decision

Overall satisfaction with decision was rated at 4.41 out of 5 (SD=0.84) in the mainstreaming group and 4.53 out of 5 (SD=0.57) in the webinar group. Satisfaction with decision shortly after their appointment did not differ significantly between groups or based on age, gender, education, race/ancestry, or diagnosis (Supplemental Table 1).

#### Patient Knowledge and Informed Choice

In total, 62% (38/61) of participants that were offered genetic testing through the mainstreaming model demonstrated adequate knowledge (score ≥75%) and were classified as informed (Table 4). Fifty nine percent (36/61) of participants received the information sheet of which 69% (25/36) were considered knowledgeable and informed. In comparison, 52% (13/25) of participants that did not receive the patient information sheet were considered knowledgeable and informed (p=0.17).

**Table 4.** Knowledge, attitude, intention to have genetic testing, and informed choice based on model of care.

| **Model** | **Knowledgeable**<br>**(≥75%)** | **Positive**<br>**Attitude**<br>**(score ≥3/5)** | **Intention**<br>**to have**<br>**genetic**<br>**testing** | **Informed**<br>**(Knowledgeable and**<br>**attitude aligned with**<br>**intent to have**<br>**genetic testing)** |
| --- | --- | --- | --- | --- |
| Webinar Model | 110/121<br>(91%) | 120/121<br>(99%) | 120/120<br>(100%) | 110/121<br>(91%) |
| Mainstreaming<br>Model | 38/61<br>(62%) | 59/61<br>(97%) | 58/61<br>(95%) | 38/61<br>(62%) |
| Mainstreaming:<br>provided<br>information<br>sheet | 25/36<br>(69%) | 35/36<br>(97%) | 34/36<br>(94%) | 25/36<br>(69%) |
| Mainstreaming:<br>not provided<br>information<br>sheet | 13/25<br>(52%) | 24/25<br>(96%) | 24/25<br>(96%) | 13/25<br>(52%) |

Significantly more webinar participants demonstrated adequate knowledge (91%; n=110/121; p < 0.01) and the same proportion were classified as having made an informed choice. Informed choice did not significantly differ based on age, gender, education, race/ancestry, or diagnosis (Supplemental Table 2).

The mean knowledge score was 77% (SD 20%) in the mainstreaming group and 89% (SD 13%) in the webinar group. The proportion of correct responses to the eight true/false statements for both groups are shown in Figure 3. Participants struggled most with the statements addressing limitations of genetic testing (Q2), followed by variable expression (Q5), and the possibility of an uncertain result (Q8).

**Figure 3:**
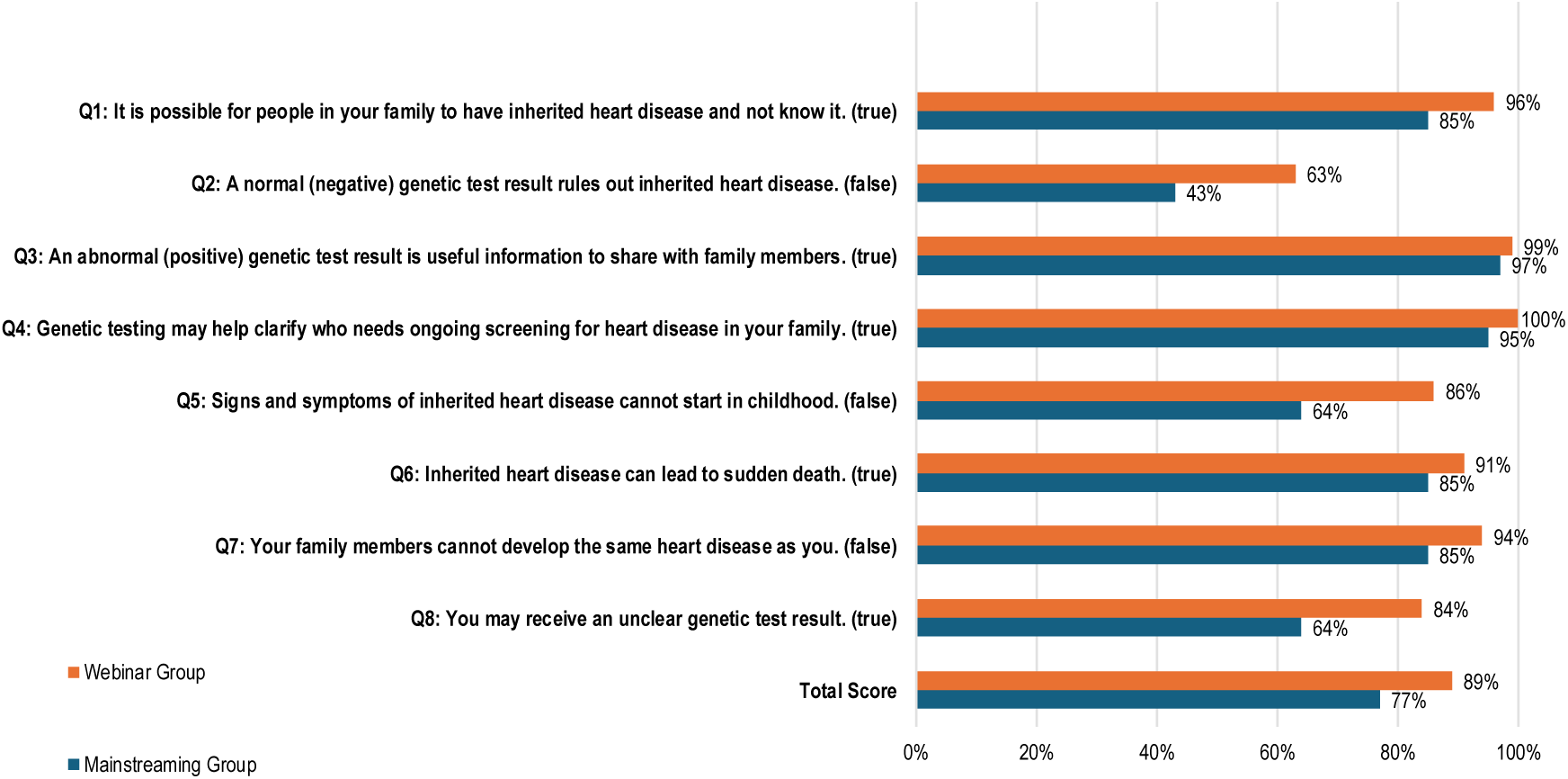
Percent of correct responses to knowledge statements in mainstreaming group compared to webinar group. Q= Question

## Discussion

This study highlights key challenges and opportunities in the evolving landscape of cardiac genetic testing. Our findings demonstrate that by mainstreaming genetic testing we were able to improve access to genetic testing and dramatically reduce wait times for patients to be offered testing. However, patients were better informed about genetic testing when offered testing by a genetic counsellor. Overall, 82% of patients with DCM or HCM that were offered cardiac genetic testing through mainstreaming had testing completed compared to 69% of patients referred to Medical Genetics. The difference in access was influenced by the immediacy of sample collection for in-patients seen through the mainstreaming model and patients that never followed through with their referral to Medical Genetics. In contrast, 91% of patients offered genetic testing by a genetic counsellor made an informed choice compared to only 62% of patients offered testing by a cardiac specialist.

The positive impacts of mainstreaming on access to genetic testing have similarly been reported in cancer genetics. For example, Chai et al. found that 88% of patients offered genetic testing through mainstreaming completed genetic testing compared to 51% of patients sent through the traditional medical genetics referral model.^21^ Numerous other studies report the advantage of reduced wait times for testing.^9^ Patient satisfaction has also previously been reported as high for patients seen through cancer mainstreaming models. More limited data is available regarding knowledge: Bokkers et. reported slightly higher knowledge scores in their control group (cancer genetics) compared to their mainstreaming group, however the difference was not significant (p=0.155).^22^ We are not aware of any studies that have evaluated informed choice as an outcome for mainstreaming of cancer genetic testing.

The findings of our study raise a conceptual question: in the context of cardiac genetics, is it more important that testing is widely accessible, or that patients are well informed? The answer likely depends on the clinical utility of genetic testing. Identifying a pathogenic variant confirms the underlying etiology and enables cascade genetic testing. This permits the identification of at-risk family members while safely discharging non-carriers from lifelong clinical surveillance. One could argue that when the value lies in predictive cascade testing, being better informed may be more important as non-informed patients may struggle in effectively communicating risk information with family members. Conversely, when clinical utility centres on guiding medical management and risk stratification, timely access becomes increasingly important. Current guidelines recommend a lower threshold for primary prevention ICD implantation in carriers of high-risk genotypes such as *LMNA, FLNC, DSP*, and *RBM20*.^23^ A definitive genetic diagnosis also unlocks targeted treatment in some cases; enzyme replacement therapy in Fabry disease being a clear example.^23^

Ideally, neither access nor informed decision making would be compromised. To address the dual goal of improving both, several strategies may be considered. Increasing the number of cardiac genetic counsellors and embedding them within cardiology clinics would ensure expert guidance at key decision points. Further, exploring acceptable and convenient alternative sample types (ie saliva or buccal swabs) for testing could further improve access across both models of care by eliminating the need for yet another appointment to have a blood sample collected.

In terms of improving understanding, an assessment of patient resources and genetics education for cardiac specialists could be considered. Cardiac specialists in this study were provided with a two-page information sheet to share with patients. We found that 69% of patients who received the education material were able to make an informed choice compared to 52% who did not. Future work could evaluate educational materials to determine whether key concepts are being effectively communicated and to identify opportunities for refinement. Additionally, the use of alternative educational modalities, such as multimedia or online tools, may further enhance patient understanding while circumventing potential time constraints faced by cardiac specialists. Notably, patient videos have been shown to be noninferior to in-person genetic counselling in the cancer genetics setting with respect to test uptake, patient knowledge, and satisfaction, suggesting a promising avenue for scalable education within cardiac genetics.^24,25^

Further, the cardiac specialists were provided with a 30-minute education session at the start of the study. Development of more extensive genetic education for cardiac specialists interested in offering genetic testing should be considered in addition to some form of genetics curriculum for cardiac fellows during their training.

Within mainstreaming models, consideration should also be given to the genetic counselling resources required for patients with positive and VUS results to address post-disclosure navigation, segregation analysis, cascade genetic testing in at-risk family members, and reproductive counselling. With increased access through mainstreaming, we may observe a shift in wait times from pre-test to post-testing counselling, highlighting the need for more innovative ways to manage these referrals.

Finally, we found that the most challenging concept for patients to understand in both groups was that a negative genetic test result does not exclude a genetic cause for their cardiac condition. Our understanding of the genetic basis of inherited cardiac conditions is still evolving as there are disease-associated genes yet to be discovered, and some genetic causes are not included on current testing panels. In addition, standard genetic testing may not detect all types of genetic variation, such as deep intronic variants, structural variants, epigenetic changes, or variants in regulatory regions that affect gene expression. This finding underscores the importance of post-test education for patients receiving a negative result, particularly for those diagnosed at a young age and/or with positive family history. In many mainstreaming models, patients with negative results will not have follow-up contact with a genetic counsellor and it is therefore key that cardiac specialists provide patients with clear guidance regarding screening recommendations for their at-risk relatives. Alternatively, patients with a negative genetic result could be referred for post-test genetic counselling leading to the need for more genetic counselling resources.

Collectively, the findings of this study underscore the need for balanced, flexible care models that recognize the variable utility of genetic testing. Future research should continue to evaluate patient outcomes, the effectiveness of integrated genetic counselling approaches, and the degree to which educational resources for patients aid in informed decision-making across diverse genetic specialties.

## Study Limitations

This study has several limitations that should be considered when interpreting the findings. First, it was conducted within a publicly funded healthcare system, where cost is not a direct barrier to access; therefore, the findings may not be generalizable to settings where financial considerations influence uptake of genetic testing. Second, we did not capture data on cardiology patients who declined genetic testing at the outset of the mainstreaming pathway, which limits our ability to definitively assess testing uptake within this group. However, there is a good chance that these individuals would have declined a referral to Medical Genetics through the traditional model and this data would also not have been captured in the referral cohort. Third, informed choice was not evaluated for patients who received genetic counselling through traditional one-on-one appointments, restricting direct comparison of informed decision-making across service delivery models. Finally, baseline genetic knowledge was not assessed prior to testing, which limits our ability to determine how pre-existing knowledge may have influenced patient understanding or decision-making following education or counselling.

It should also be noted that all cardiac specialists in this study practiced, at least in part, within a tertiary care setting, and most had prior familiarity with genetic testing before participation in the study. The study allowed the cardiac specialists to provide genetic testing for the disorders they subspecialize in and the specialists were supported by Medical Genetics infrastructure for post-test counselling and follow-up. As such, these findings may not fully reflect the experiences or needs of cardiac specialists with less familiarity of the clinical utility of genetic testing and confidence in interpreting results.^26,27^ This highlights the importance of accessible, targeted educational resources to support the successful implementation of mainstreaming models across diverse practice environments. While some educational tools currently exist, further development and evaluation of scalable, specialty-specific resources will be essential to support informed test ordering, result interpretation, and appropriate patient counselling.

## Conclusion

In conclusion, integrating genetic testing directly into cardiology care through a mainstreaming model improves access and significantly reduces pre-test wait times, allowing patients with HCM and DCM to move forward with testing sooner than through traditional referrals. However, while this approach enhances efficiency, the findings show that patients in the mainstreaming pathway had substantially lower knowledge and rates of informed choice compared with those seen through a structured webinar model. This highlights the need for additional support, such as standardized educational materials, decision aids, and reliable referral pathways to genetic counsellors, to optimize the patient experience within mainstreaming. Ultimately, a hybrid model in which cardiologists initiate testing while genetics professionals provide targeted support, particularly for complex decisions or positive findings, may offer the best balance between timely access and comprehensive, informed patient care.

## Data Availability

The data is available upon request.

## Ethics Statement

This article adheres to the relevant ethical guidelines

## Patient Consent Statement

The authors confirm that patient consent forms have been obtained for this article.

---

**Supplementary Table 1:** Decisional Satisfaction based on demographic variables.

| Demographic Variable | Decisional Satisfaction: Medical Genetics Webinar |  | Decisional Satisfaction: Mainstreaming |  |
| --- | --- | --- | --- | --- |
|  | Coefficient [95% CI] | p value | Coefficient [95% CI] | p value |
| HCM vs DCM | 0.04 [-0.18, 0.26] | 0.72 | 0.43 [-0.71, 1.57] | 0.45 |
| Female vs male | 0.08 [-0.13, 0.30] | 0.42 | -0.40 [-1.49, 0.70] | 0.47 |
| ≥50 years vs <50 years | -0.05 [0.26, 0.16] | 0.62 | -0.60 [-1.69, 0.49] | 0.28 |
| Post high school education vs no post high school education | 0.12 [-0.15, 0.19] | 0.85 | -0.58 [-1.70, 0.54] | 0.30 |
| Non-White vs White Ethnicity | -0.01 [-0.23, 0.22] | 0.95 | -0.17 [-1.46, 1.12] | 0.80 |

**Supplementary Table 2:** Informed choice based on demographic variables.

| Demographic Variable | Informed choice: Medical Genetics Webinar |  | Informed choice: Mainstreaming |  |
| --- | --- | --- | --- | --- |
|  | Odds Ratio [95% CI] | p value | Odds Ratio [95% CI] | p value |
| HCM vs DCM | 0.78 [0.21, 2.93] | 0.71 | 0.98 [0.32, 2.97] | 0.97 |
| Female vs male | 0.50 [0.14, 1.75] | 0.28 | 0.94 [0.32, 2.75] | 0.91 |
| ≥50 years vs <50 years | 0.91 [0.26, 3.17] | 0.89 | 0.82 [0.28, 2.37] | 0.71 |
| Post high school education vs no post high school education | 1.93 [0.57, 6.60] | 0.29 | 0.91 [0.31, 2.67] | 0.86 |
| Non-White vs White Ethnicity | 0.55 [0.19, 1.55] | 0.26 | 0.47 [0.14, 1.59] | 0.22 |

